# Assessing the effectiveness of online emotion recognition training in healthy volunteers

**DOI:** 10.1101/2023.03.10.23286897

**Authors:** Zoe E Reed, Steph Suddell, Andy Eastwood, Lilian Thomas, Imogen Dwyer, Ian S Penton-Voak, Christopher Jarrold, Marcus R Munafò, Angela S Attwood

## Abstract

**Background:** Difficulties in facial emotion recognition are associated with a range of mental health and neurodevelopmental conditions and can negatively impact longer term social functioning. Interventions that target facial emotion recognition may therefore have important clinical potential, for example for autistic individuals. We investigated the effect of an emotion recognition training (ERT) task on emotion recognition ability and, importantly, whether training generalises to novel (non-trained) faces.

**Methods:** We conducted three online experimental studies with healthy volunteers completing a single ERT session to test: 1) the efficacy of our four-emotion ERT (training to improve recognition of angry, happy, sad and scared emotional expressions) (N=101), 2) the efficacy of our six-emotion ERT (adding disgusted and surprised) (N=109), and 3) the generalisability of ERT to novel (non-trained) facial stimuli (N=120). In all three studies, our primary outcome was total correct hits across all emotions. In Studies 1 and 2, this was compared across active training and control (sham) training groups (randomised). In Study 3, this was compared across groups who were trained on stimuli that were either the same identity (stimulus-congruent), or a different identity (stimulus-incongruent) to those they were tested on (randomised). Linear mixed effects models were used to test for effects of training.

**Results:** The effect estimate from Study 1 was in the direction of improvement in the active training group, however, confidence intervals were wide (*b*=0.02, 95% CI=-0.02 to 0.07, *p*=0.27) and our effect may have been reduced due to ceiling effects. Study 2, with the additional emotions, indicated total hits were greater following active (vs. sham) training, which remained following inclusion of baseline covariates (*b*=0.07, 95% CI=0.03 to 0.12, *p*=0.002). Study 3 demonstrated that improvement post-training was similar across stimulus-congruent and incongruent groups (*b*=-0.01, 95% CI=-0.05 to 0.02, *p* = 0.52).

**Conclusion:** Our results indicate that ERT improves emotion recognition and that this improvement generalises to novel stimuli. Our data suggest six emotions should be used rather than four to avoid ceiling effects in training. Future studies should explore generalisability of facial stimuli of different ages and ethnicities as well as examining longer-term effects of ERT. The application of ERT as an intervention may be particularly beneficial to populations with known emotion recognition difficulties, such as autistic individuals.

## 1. Introduction

The ability to perceive and recognise facial emotional expressions is an important facet of social cognition (Beaudoin & Beauchamp, 2020) and an essential non-verbal tool for interpersonal communication – enabling us to infer the mental states of others (Mier et al., 2010). Human social behaviour, such as engaging with or avoiding other people, is also influenced by the perception of emotions (Seidel et al., 2010). Previous research has suggested that difficulties in this sociocognitive domain (i.e., recognising others’ emotions) is associated with a range of mental health and neurodevelopmental conditions, greater internalising and externalising behaviours, and problems in areas such as social competence, academic ability and social skills (Denham et al., 2015; Izard et al., 2001; Leppänen & Hietanen, 2001; Trentacosta & Fine, 2010). For example, poorer emotion recognition has been found in individuals with depression (Dalili et al., 2015; Demenescu et al., 2010), and this may be a causal factor in the maintenance of depressive symptoms (Warren et al., 2015). Similarly, autistic individuals or those scoring higher on autistic trait measures tend to have lower accuracy for global emotion recognition (Law Smith et al., 2010; Lozier et al., 2014; Reed et al., 2021). This may have implications for social development and interpersonal skills over time (Uljarevic & Hamilton, 2013). Developing a training paradigm to improve emotion recognition accuracy may have therapeutic benefits for a number of mental health and neurodevelopmental conditions (Penton-Voak et al., 2017). However, given that autistic individuals, in particular, seem to experience difficulties with global emotion recognition (as opposed to specific emotional biases), then developing an intervention for emotion recognition, targeting autistic individuals specifically, is important. In fact, previous research has highlighted the potential for such interventions in autism, but many lack a strong evidence base and it is unclear whether training gains generalise beyond trained stimuli (Kouo & Egel, 2016).

Previous research has shown that is possible to attenuate atypical emotion recognition *biases* (i.e., a tendency to consistently interpret ambiguous facial expressions as a particular emotion), which are present in several mental health conditions, using a brief digital training task. For example, biases have been found towards perceiving sadness in anxiety and depression, and anger or disgust in alcohol use disorder (Freeman et al., 2018; Penton-Voak et al., 2017). Training studies in anxious and depressed samples have shown decreased biases to negative emotions post-training, and that this decrease is associated with some improvement in mood and quality of life, although these therapeutic benefits are inconsistent across studies (Penton-Voak et al., 2012; Suddell et al., 2021).

Using a similar approach, we have developed a computer-based task that measures the ability to *recognise* emotional expressions in faces (i.e., identifying the correct emotion for a facial expression) (Griffiths et al., 2019). However, it remains unclear whether this emotion recognition task can be adapted to *improve* recognition of emotional expressions as well as measure them, similar to the tasks previously developed that can modify biases in emotional expressions. Here, we assess whether our adapted training task can improve emotion recognition. This initial evidence would be an important step towards the development of emotion recognition interventions, that may be beneficial for autistic individuals, and others, that have difficulties in this area.

When developing a training paradigm, as well as improving the outcome of interest, it is essential that effects generalise to wider contexts outside of the specific study conditions. The majority of emotion recognition research uses static facial stimuli (de Paiva-Silva et al., 2016), with some research demonstrating that emotion recognition training (ERT) can generalise outside of the initial training setting and to different stimuli (Dalili et al., 2016; Griffiths et al., 2015). Therefore, it would be useful to also assess if that is the case for our ERT.

Here we investigate whether an ERT task we have developed improves emotion recognition accuracy. We conducted three online experimental studies. The protocols for each study were pre-registered on the Open Science Framework. Study 1 (https://osf.io/x4kh3) aimed to test the effect of a four-emotions (angry, happy, sad and scared) version of our ERT task. Study 2 (https://osf.io/drby2) tested the same task with six emotions (additional emotions of disgusted and surprised). Study 3 (https://osf.io/bpzcj) tested whether training effects generalised to novel (non-trained) faces. We hypothesised that there would be an increase in emotion recognition accuracy (measured by total hits) after active ERT compared to sham training (Studies 1 and 2). We also hypothesised that training effects would be present and comparable between two groups trained on previously seen (congruent) versus two groups trained on previously unseen (incongruent) stimuli (i.e., that effects would generalise to non-trained faces).

## 2. Study 1

### 2.1 Methods

For all studies, healthy volunteers were recruited through the online recruitment platform Prolific (https://www.prolific.co/) and data were collected via Gorilla, an online experiment builder (http://www.gorilla.sc/) (Anwyl-Irvine et al., 2019). All studies consisted of a single session of ERT. Ethics approval was obtained from the School of Psychological Science Research Ethics Committee at the University of Bristol for all studies.

Data and code availability: The data and analysis code that form the basis of the results presented here for all studies are available from the University of Bristol’s Research Data Repository (http://data.bris.ac.uk/data/), DOI: https://doi.org/10.5523/bris.1df0stlnxblc72a13mfnsne3ew.

#### 2.1.1 Participants

We recruited 110 healthy volunteers randomised in a 1:1 ratio to one of two training groups (active or sham). To participate in this study, participants had to be aged 18 or over and fluent in English. Exclusion criteria included ever being diagnosed with any mental health condition, currently using psychiatric medication, and having an uncorrected visual impairment (including colour blindness). Screening was based on self-report within the participants’ Prolific profiles and screening questions were also asked within the study to verify eligibility.

Sample size was determined based on a previous study reporting an effect size of *d*=1.08 for the effect of emotional *bias* training on the perception of happy faces (Penton-Voak et al., 2012). We used a more conservative effect size of *d*=0.70 to take account of the possibility that the original observed effect size may have been inflated as initial studies tend to report inflated effects if evidence of an effect is based on crossing a threshold of statistical significance (e.g., p<0.05) (Ioannidis, 2008). We calculated that, at an alpha level of 5% for a two-tailed independent means t-test, 110 participants would provide 95% power to detect an effect size of *d*=0.70, 90% power to detect an effect size of *d*=0.63 and 80% power to detect an effect size of *d*=0.54.

#### 2.1.2 Study procedure

Demographic information on age and gender were collected. All participants completed an initial baseline emotion recognition four alternative forced choice (4AFC) test, measuring recognition of anger, happiness, sadness and fear in male facial stimuli. Participants in the active group then completed a similar emotion recognition 4AFC training task in which they received feedback as to whether they had responded correctly or not. If they responded incorrectly, they had to keep responding until they selected the correct emotion.

Participants in the sham training group completed a 4AFC training task with feedback, but the stimuli consisted of boxes displaying different colours rather than facial stimuli and participants were asked to select which colour they thought was being displayed. After completing their respective training tasks, participants completed a final emotion recognition 4AFC test and questions on subjective outcomes and the positive and negative affect schedule (PANAS).

#### 2.1.3 Emotion recognition test: baseline and post-training

The task used as the baseline and post-training emotion recognition tests presented four facial emotional expressions (angry, happy, sad and scared). There were 15 levels of intensity presented for each emotion, resulting in a total of 60 trials. Images of facial expressions were shown for 150 milliseconds (ms) with backward masking. On the next screen participants were asked to select the descriptor that best described the emotion displayed and there was no time limit for this. Presentation of the facial stimuli was randomised. Images used were of the same (male) individual. Figure 1 displays a trial schematic of the emotion recognition test.

**Figure 1.**
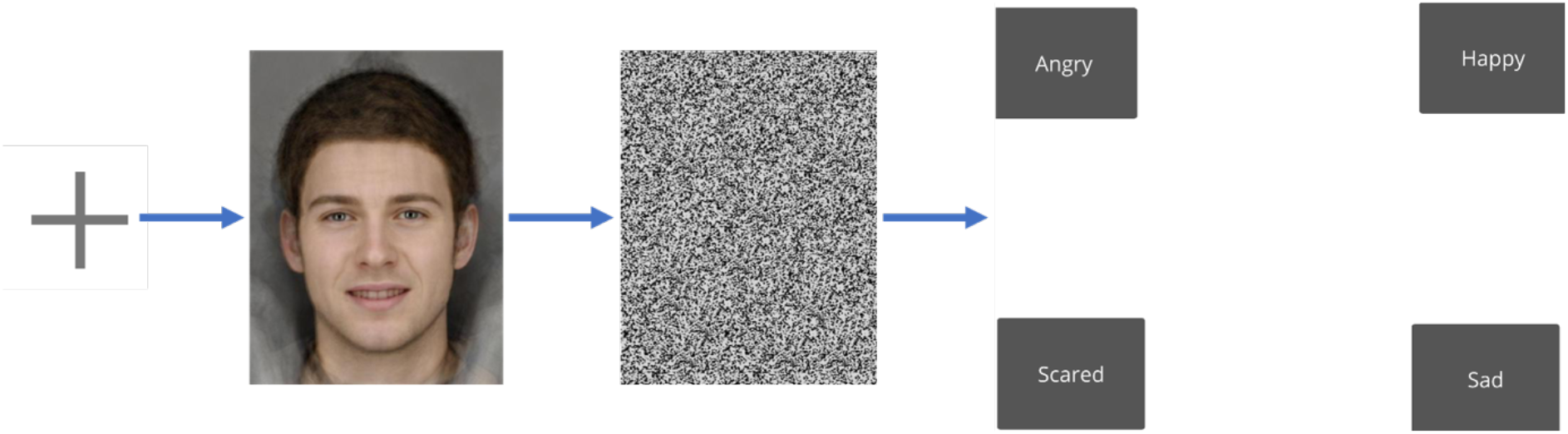
Example of emotion recognition task with happy emotion facial stimuli: First a fixed cross is shown for 800ms, then an image of a face displaying an emotion for 150 milliseconds, which was backwards masked for 250ms and finally a screen with the four emotions (angry, happy, sad and scared) is shown. The participant selects an emotion as their response. Facial stimuli are computer generated by averaging photos of 12-15 individuals and therefore do not show a real person.

#### 2.1.4 Active emotion recognition training (ERT) task

The active training group completed a training version of the emotion recognition 4AFC test described above. The procedure was the same as for the test, except that the face was shown for one second and response feedback was provided. The participant had to keep responding until they answered correctly.

#### 2.1.5 Sham (colour) training task

The sham training group completed a similar training task to the active group, again with response feedback. However, the stimulus set consisted of coloured blocks instead of facial stimuli. The colours used for these blocks were blue, red, green and yellow for the 4AFC task. These colours were presented at the same number of levels of intensity as facial stimuli, on a greyscale (i.e., ranging from grey through to the respective colour in 15 increments) and participants were asked to select which colour they thought was presented.

#### 2.1.6 Subjective outcomes

Participants were asked the following additional questions related to the study as a whole: “Did you find the task fatiguing?”, “Did you find the task interesting” and “did you find the task challenging?”. Participants were asked to provide responses on a scale of zero to 100, where zero indicated “not at all” and 100 indicated “very much so”.

In addition, participants were asked to provide responses to 20 items from the PANAS (Watson et al., 1988). These items were “interested”, “distressed”, “excited”, “upset”, “strong”, “guilty”, “scared”, “hostile”, “enthusiastic” “proud”, “irritable”. “alert”, “ashamed”, “inspired”, “nervous”, “determined”, “attentive”, “jittery”, “active” and “afraid”. Participants indicated the extent to which they currently felt each of the emotions/feelings from the options “very slightly or not at all”. “a little”, “moderately”, “quite a bit”, “extremely”.

#### 2.1.7 Outcome measures

For statistical analyses our outcomes included: 1) total hits in the baseline and post-training tasks, i.e., the total number of correct responses (indicative of emotion recognition accuracy), 2) hits per emotion, 3) false alarms per emotion (i.e., the number of times a particular emotion was selected when this was not the correct response), 4) sensitivity scores per emotion using the signal detection theory Aprime (A’) index which is a non-parametric estimate of discriminability. Sensitivity scores were calculated using the Dprime function from the R ‘Psycho’ package (Makowski, 2018) which calculates a number of indices. We used the A’ measure, which is a non-parametric estimate of discriminability, where values near 1 indicate good discriminability and values near 0.5 indicate poorer discriminability (i.e., closer to chance). The R function uses the Hautus adjustment (adding 0.5 when calculating A’ so that where participants have 1 for hit rate or 0 for false alarm rate the A’ can still be calculated). Hits and false alarm outcomes were converted to proportions for analyses.

After removal of outliers, skewness and kurtosis measures were examined. Histograms of the distribution of total hits as a proportion at baseline and post-training are shown in Supplementary Figure S1. Skewness and kurtosis measures were within an acceptable range (see Supplementary Materials, Section 2, for further detail).

#### 2.1.8 Statistical analysis

All analyses were conducted in R version 4.0.0 or 4.0.2 depending on when analyses were conducted (R Core Team, 2020). We compared group differences (active versus sham training) using a linear mixed effects (LME) model with total hits as the outcome and time, group and an interaction term for time x group as the predictors, while accounting for between subjects (specified as participant ID) random variance. To do this, we used the lme4 package in R (Bates et al., 2015), where fixed effects of time and group were included in the LME model. Finally, random intercepts for each participant were included for the random effects. The LME model is different to the planned ANOVAs included in our pre-registered protocol. We made this change to the analysis plan as this model allows for more control over random (e.g., participant ID) and fixed (i.e., group, time, an interaction term and covariates) factors. It also allowed us to model all data points rather than aggregated data as in an ANOVA.

We conducted secondary analyses using LME models to explore hits, false alarm rates and sensitivity scores across the individual emotions with the same predictors as in the primary outcome model. Finally, we assessed whether each of the subjective ratings of training experience and the PANAS positive and negative scores (from adding together individuals item scores) varied between the two groups by conducting two-tailed independent means t-tests. The active and sham tasks were different, so we examined subjective ratings of the tasks to see how participants found them across the groups.

## 2.2 Results

### 2.2.1 Exclusion of participants

We recruited 110 participants. However, after removing those from analyses that did not meet eligibility criteria (N=5), or whose data were outliers for total hits (below 0.60 and 0.72 for baseline and post-training hits, respectively) (N=4), there were 101 participants included in our analyses (52 in active and 49 in sham groups).

### 2.2.2 Participant characteristics

Table 1 shows the participant characteristics.

**Table 1.**
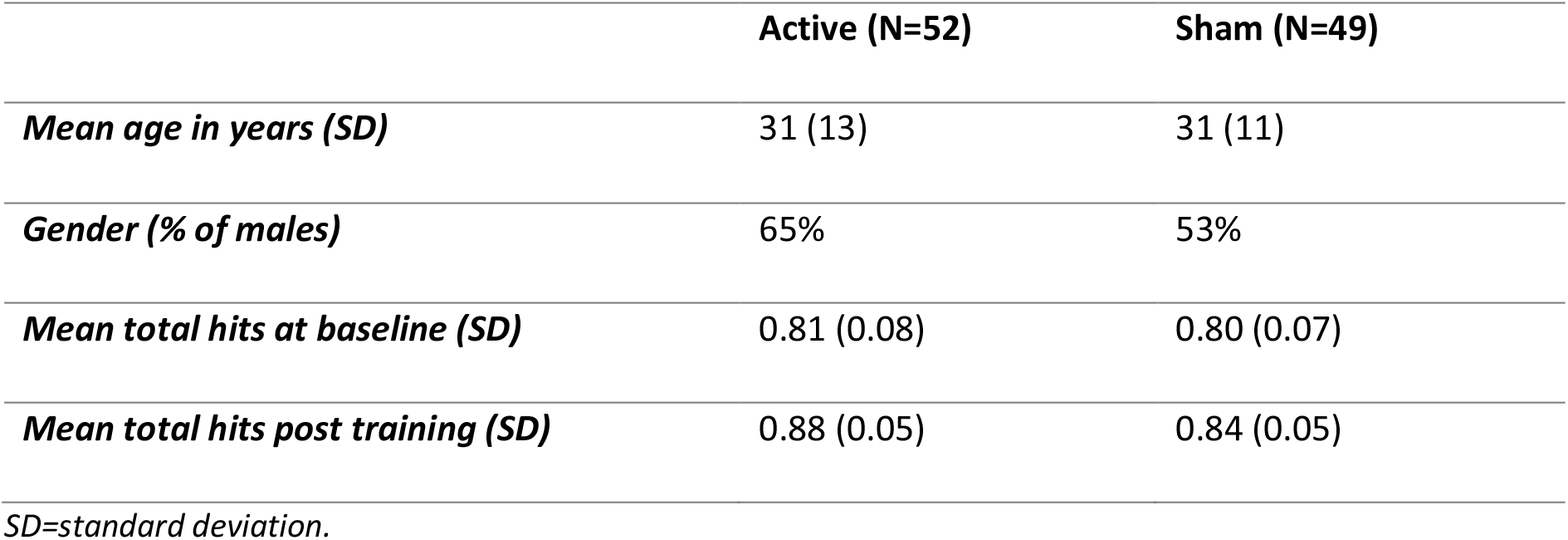
Study 1 sample descriptives

### 3.2.3 Analysis results

We found that total hits were greater post-training, with a 6% increase in recognition accuracy in our main effects model (*b*=0.06, 95% CI=0.04 to 0.08, *p*<0.001) and greater in the active group (*b*=0.02, 95% CI=0.002 to 0.05, *p*=0.03) in our main effects model, but there was no clear statistical evidence for the effect of training condition over time in our interaction model (*b*=0.02, 95% CI=-0.02 to 0.07, *p*=0.27) (Figure 2 and Supplementary Table S1).

**Figure 2.**
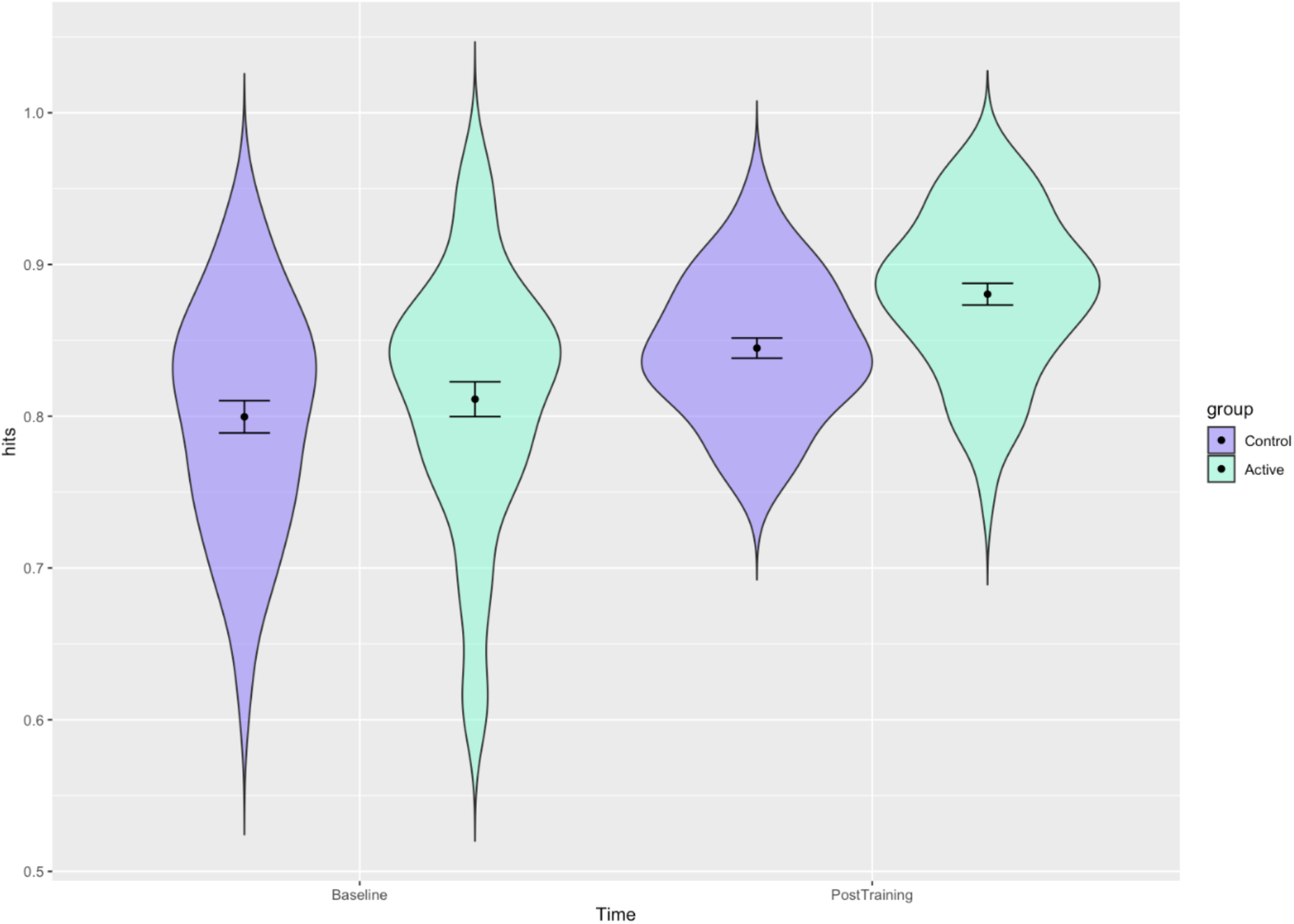
Distribution of participant’s total hits with estimates for the active and sham groups at baseline and post training in Study 1. Error bars represent 95% confidence intervals: Distribution of participants scores (in terms of proportion of total correct hits) with estimates for each group before and after training and confidence intervals shown. The active group shows a slight improvement post-training.

Results from the LME models for emotion specific hits, false alarms and sensitivity scores are presented in Supplementary Tables S2-4. We found little evidence for an interaction effect between time and group on the number of hits for most emotions in the post-training active sham groups compared to pre-training, except for anger where there was an increase (*b*=0.07, 95% CI=0.01 to 0.13, *p*=0.03). Here, our estimate at baseline for the sham group (i.e., the intercept) is 64% and it is 67% for the active group. After training this increased to 73% in the sham group and 83% in the active group. For false alarms there was also little change, apart from a decrease in happy false alarms in the interaction model (*b*=-0.04, 95% CI=-0.07 to -0.01, *p*=0.02), indicating fewer false alarms. For the sham group at baseline this was 10% and for the active group this was 9%, decreasing to 9% post training for the sham group and 4% for the active group. There was little evidence for an interaction effect between time and group on sensitivity scores in the post-training active sham groups compared to pre-training, although we did find weak evidence for an increase for happy (*b*=0.02, 95% CI=-0.001 to 0.04, *p*=0.07), where the sham group increased from 0.93 to 0.94 and the active group increased from 0.92 to 0.95, which was likely driven by the decrease in false alarms.

Finally, we did not find any differences between the two groups for most of the subjective ratings or the PANAS negative scores (Supplementary Table S5). There was weak evidence of a difference for the challenging subjective rating, with the active group reporting that this was more challenging than the sham group (active mean=63 [SD=23], sham mean=54 [SD=28], *p*=0.09). The active group also reported slightly lower positive scores than the sham group for the PANAS (active mean=27 [SD=8], sham mean=30 [SD=8], *p*=0.07).

## 3. Study 2

### 3.1 Methods

#### 3.1.1 Participants

We recruited 116 healthy volunteers randomised in a 1:1 ratio to one of two groups (active or sham). Exclusion/inclusion criteria was applied as described in Study 1, with the additional exclusion criterion of having participated in Study 1.

We used the same power calculation to determine the sample size as in Study 1 but with an additional increase to participant numbers of 5% based on having to exclude this percentage of participants in Study 1 due to them not meeting the screening criteria within the study.

#### 3.1.2 Study procedure

The procedure was similar to that of Study 1 except with a six alternative forced choice (6AFC) design, which additionally included disgusted and surprised emotional expressions in male facial stimuli. We also collected additional information on education at baseline and questions from the 10-item Autism Spectrum Quotient (AQ-10) (Allison et al., 2012; Booth et al., 2013) at the end of the session instead of the PANAS used in Study 1.

#### 3.1.3 Emotion recognition test: baseline and post-training

Similar to Study 1, in Study 2 the ER test was used at baseline and post-training. However, in addition to the four emotions used in Study 1, Study 2 included two additional emotions of disgust and surprise. Also, to reduce the number of trials due to the increased number of emotions/colours, participants were only presented with 8 levels of intensity for each stimulus (every other level of intensity of the original 15), resulting in a total of 48 trials. The task was the same in all other respects to that used in Study 1.

#### 3.1.4 Training tasks

The active training group completed the same task as described in Study 1 with the addition of 2 emotions as described for the test above.

The sham training group completed the same task as described in Study 1 as well, with the addition of orange and purple coloured blocks for the 6AFC task with 8 increments.

#### 3.1.5 Subjective outcomes

Participants were asked the following additional questions related to the study as a whole: “Did you find the task tiring”, “Did you find the task interesting” and “did you find the task challenging?”. Participants were asked to provide responses on a scale of zero to 100, where zero indicated “not at all” and 100 indicated “very much so”. In addition, participants were asked to provide responses to the AQ-10 (Supplementary Materials Section 1).

#### 3.1.6 Outcome measures

For statistical analyses our outcomes were the same as those for Study 1: 1) total hits in the baseline and post-training tasks, 2) hits per emotion, 3) false alarms per emotion, and 4) sensitivity scores per emotion. Histograms of the distribution of total hits as a proportion at baseline and post-training are shown in Supplementary Figure S2. Skewness and kurtosis measures were within an acceptable range (see Supplementary Materials, Section 2, for further detail).

#### 3.1.7 Statistical analysis

Similar, to Study 1 we compared group differences (active versus sham training) using an LME model with the same predictors. Here, we ran models both excluding and including covariates (for age, gender, education level (as fixed effects)), to improve precision of our effect estimate as these variables are likely to influence our outcome.

We conducted secondary analyses to assess whether the total score on the AQ-10 had any effect on total hits. To do this we ran a final model including age, gender, education level and the total score on the AQ-10. We ran LME models for each emotion for hits, false alarms and sensitivity scores as exploratory outcomes with the same covariates as in the primary outcome model. Finally, we assessed whether each of the subjective ratings of training experience varied between the two groups by conducting two-tailed independent means t-tests.

## 3.2 Results

### 3.2.1 Exclusion of participants

We recruited 116 participants. However, after removing those from analyses that did not meet eligibility criteria (N=4), or whose data were outliers for total hits (below 0.43 and 0.37 for baseline and post-training hits, respectively) (N=2), there were 109 participants included in our analyses (54 in active and 55 in sham groups).

### 3.2.2 Participant characteristics

Table 2 shows the participant characteristics.

**Table 2.** Study 2 sample descriptives

|  |  | Active (N=54) | Sham (N=55) |
| --- | --- | --- | --- |
| **Mean age in years (SD)** |  | 28 (10) | 29 (10) |
| **Gender (% of males)** |  | 67% | 67% |
| **Education (%)** | *Degree or equivalent* | 54% | 71% |
|  | *A-levels of equivalent<sup>1</sup>* | 28% | 22% |
|  | *GCSEs (grades A\* to C) or equivalent<sup>2</sup>* | 6% | 4% |
|  | *None or unsure* | 13% | 4% |
| **Mean total hits at baseline (SD)** |  | 0.64 (0.08) | 0.64 (0.09) |
| **Mean total hits post training (SD)** |  | 0.75 (0.10) | 0.67 (0.10) |

*SD=standard deviation. <sup>1</sup>A-levels or Advanced level qualifications are subject specific qualifications in the UK that are typically completed over 2 years between the ages of 16 and 18 (although can be completed over different time periods and different ages), <sup>2</sup>GCSEs (General Certificate of Secondary Education) are subject specific qualifications in the UK that are typically completed over 3 years towards the end of secondary school education. A\* was the highest possible grade.*

### 3.2.3 Analysis results

We found that total hits were greater post-training in the active group compared to the sham group in the interaction models excluding and including covariates (including covariates: *b*=0.07, 95% CI=0.03 to 0.12, *p*=0.002) (Figure 3 and Supplementary Table S6). Here the sham group hits increased from 70% at baseline to 73% post training and the active group increased from 71% to 81%. This was unchanged when additionally including AQ-10 scores (*b*=0.07, 95% CI=0.03 to 0.12, *p*=0.002).

**Figure 3.**
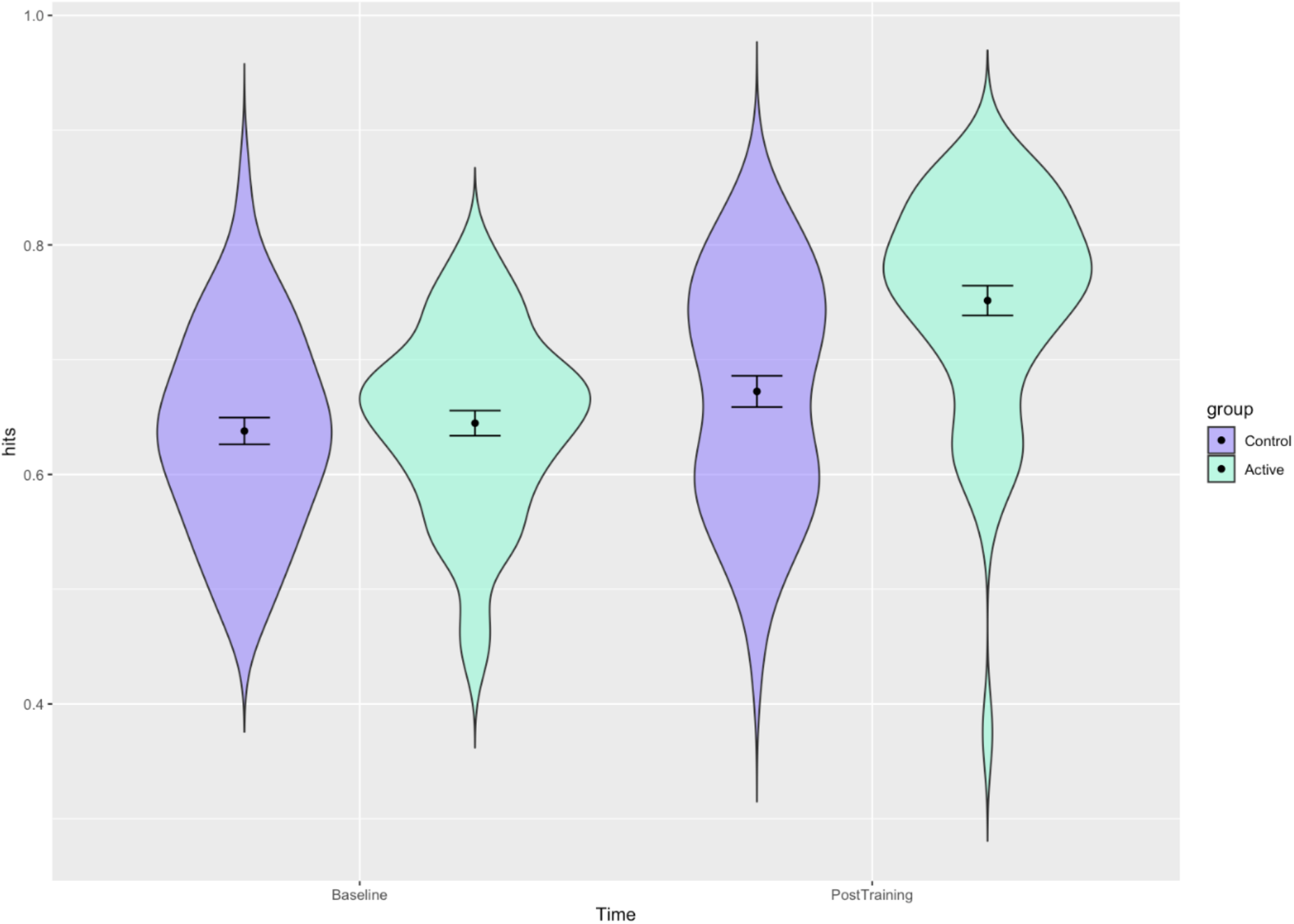
Distribution of participants’ total hits with estimates for the active and sham groups at baseline and post training in Study 2 (model including covariates). Error bars represent 95% confidence intervals: Distribution of participants scores (in terms of proportion of total correct hits) with estimates for each group before and after training and confidence intervals shown. The active group shows greater improvement post-training.

Results from LME models for emotion specific hits, false alarms and sensitivity scores including covariates are presented in Supplementary Tables S7-9. We did not find evidence for an interaction effect between time and group on the number of hits for most emotions in the post-training active sham groups compared to pre-training. However, there was some evidence for increased hits in the active group post-training for scared (*b*=0.26, 95% CI=0.17 to 0.36, *p*<.001), with the sham group increasing from 64% to 67% and the active from 65% to 94% and sad (*b*=0.07, 95% CI=-0.0002 to 0.13, *p*=0.05), with the sham group decreasing from 76% to 75% and the active group increasing from 72% to 78%. We also found decreased false alarms for surprised in the interaction model (*b*=-0.05, 95% CI=-0.07 to - 0.02, *p* < .001), with the sham group decreasing from 13% to 11% and the active group decreasing from 12% to 5% and sad (*b*=-0.02, 95% CI=-0.04 to 0.002, *p*=0.07), with the sham group decreasing from 7% to 5% and the active from 8% to 4%. There was no clear evidence for changes in the recognition of other emotions. For sensitivity scores we found some improvement post-training for the active group compared to the sham group for sad in the interaction model (*b*=0.04, 95% CI=0.01 to 0.05, *p*=0.01), with the sham group increasing from 91% to 92% and the active increasing from 89% to 94% scared (*b*=0.13, 95% CI=0.05 to 0.20, *p*=0.001), and (to a lesser extent) surprised (*b*=0.02, 95% CI=0.003 to 0.05, *p*=0.09), with the sham group increasing from 88% to 89% and the active from 87% to 90%, but no clear differences were observed for other emotions.

Finally, we did not find any meaningful differences between any of the subjective ratings for the two groups (Supplementary Table S10).

## 4. Study 3

### 4.1 Methods

#### 4.1.1 Participants

We recruited 136 healthy volunteers who were randomised to one of four training groups in a 1:1:1:1 ratio. Two of these groups were trained and tested on congruent stimuli (i.e., the set of faces were the same at both training and test, one group for male facial stimuli and the other for female facial stimuli), while the two remaining groups were trained and tested on incongruent (i.e., different) stimuli (one group tested on male and trained on female stimuli and the other tested in female and trained on male stimuli). The same exclusion/inclusion were applied as in Study 1, with the additional exclusion criterion of having participated in Study 1 or 2.

Sample size was determined through an *a priori* power calculation based on Study 2 results (*d*=1.1). Based on Study 2 being an initial trial of ERT, a more conservative effect size was used (*d*=0.70). We calculated that, with an alpha level of 5%, 120 participants would provide 95% power to detect an effect size of *d*=0.70. This was determined for the within-group comparison to assess whether training between the baseline and the post-training test was successful. We recruited an extra 8 participants per group (congruent/incongruent) to account for needing to exclude participants and having incomplete data sets. This resulted in a total of 136 participants being recruited.

#### 4.1.2 Study procedure

Study 3 used the same training and test tasks as Study 2 (6AFC), however, there was no sham condition and female faces were used in addition to male faces. Instead, all participants received active training on either congruent or incongruent facial stimuli. Participants in the congruent condition were shown the same facial stimuli (e.g., all male or all female faces) in the baseline test, training task, and post-training test. Participants in the incongruent condition were shown the same facial stimuli (e.g., male or female faces) in the baseline and post-training tests, but were presented with different stimuli (e.g., male faces if already shown female faces or female faces if already shown male faces) during training. There were four possible training conditions: male congruent, female congruent, male incongruent, female incongruent (see Figure 4). All four conditions were included to account for any asymmetrical training effects (i.e., if training on male faces transferred to female faces, but not vice versa). However, we were interested in the comparison of congruent versus incongruent and therefore for analyses we collapse these into just two groups representing these two conditions. We also collected responses on Beck’s Depression Inventory-II (BDI) scale (Beck et al., 1996). This was included for a student project, but data are not reported here as they were not part of our planned analyses. However, we included an attention check item in the BDI, stating: “this is to check you are paying attention, please select option 2”. Participants were excluded from analysis if they failed this attention check.

**Figure 4.**
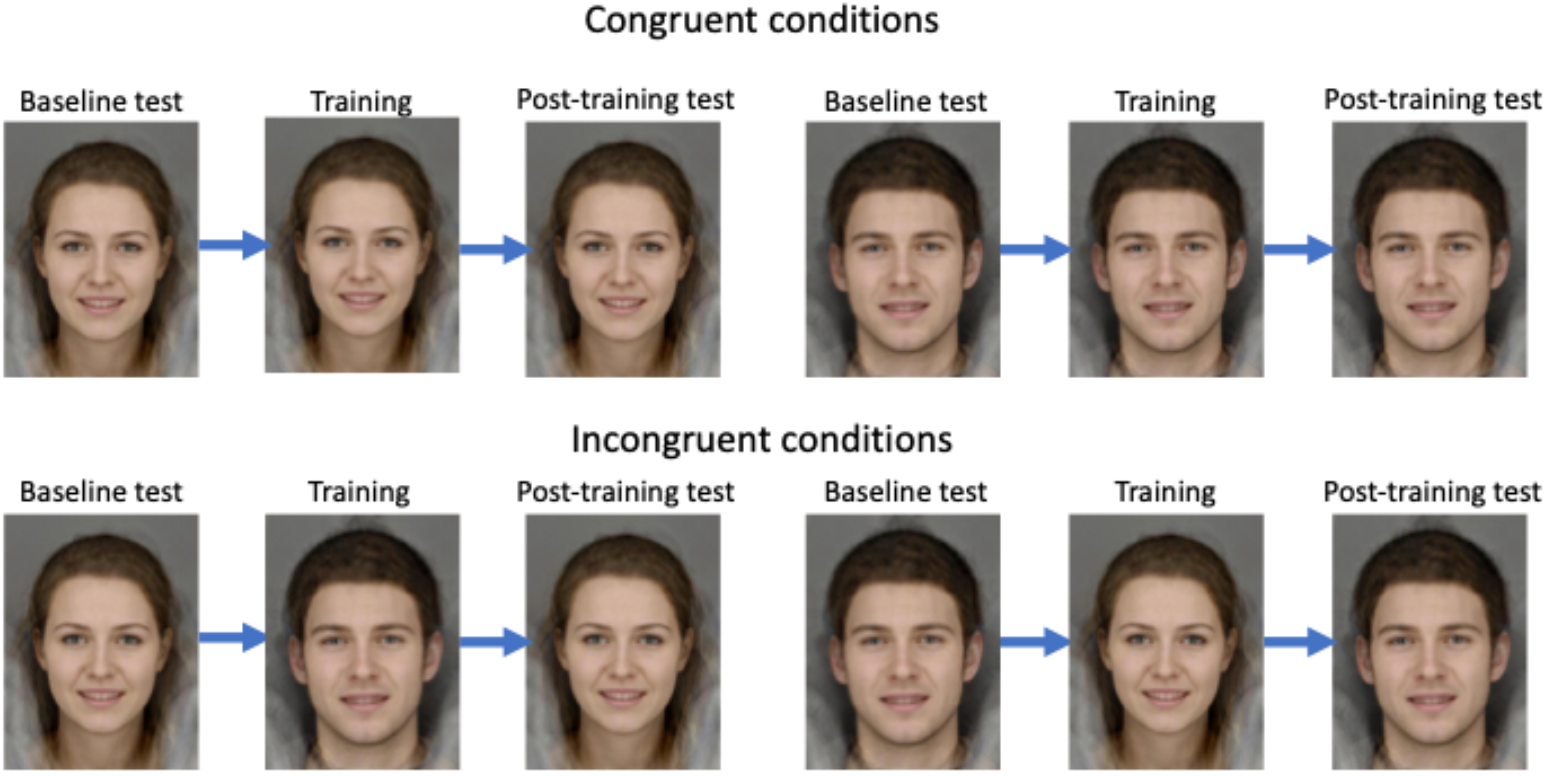
Study 3 training conditions: All four possible conditions are shown. Top-left: female congruent stimuli. Top-right: male congruent stimuli. Bottom-left and bottom-right: incongruent stimuli (either female-male-female or male-female-male). Facial stimuli are computer generated by averaging photos of 12-15 individuals per set of stimuli and therefore do not show real people.

#### 4.1.3 Outcome measures

For statistical analyses our outcome was total hits in the baseline and post-training tasks. Histograms of the distribution of total hits as a proportion at baseline and post-training are shown in Supplementary Figure S3. Skewness and kurtosis measures were within an acceptable range (see Supplementary Materials, Section 2, for further detail).

#### 4.1.4 Statistical analysis

We used an LME model to assess the effects of time (baseline vs. post-training) by stimulus congruency (congruent vs. incongruent) on proportion of total hits. We ran models without and with covariates for age, participant gender, and highest education level. The LME model is different to the planned ANOVAs included in our pre-registered protocol for the same reason as the change for Study 1, and to be consistent across studies.

## 4.2 Results

### 4.2.1 Exclusion of participants

We recruited 136 participants. However, after removing those from analyses whose data were outliers for total hits (below 0.31 and 0.50 for baseline and post-training hits, respectively) (N=5), or who failed an attention check (N=11), there were 120 participants included in our analyses (62 in congruent and 58 in incongruent groups).

### 4.2.2 Participant characteristics

Table 3 shows the participant characteristics.

**Table 3.**
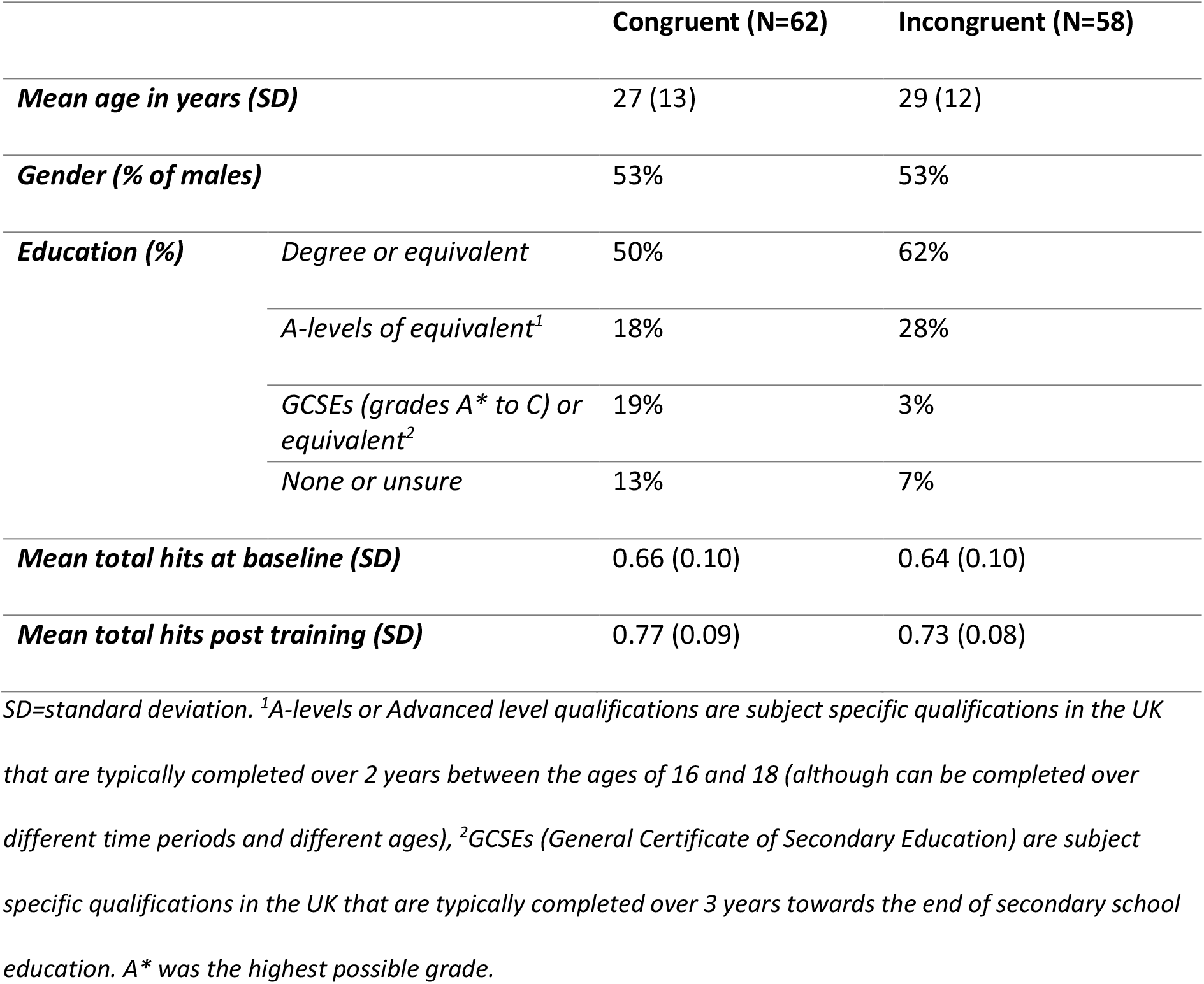
Study 3 sample descriptives

### 4.2.3 Analysis results

The LME model for proportion of total hits, including covariates, suggested an effect of time in the main effects model (*b*=0.10, 95% CI=0.08 to 0.12, *p*<.001). However, there was no evidence for an effect of stimulus congruency in the main effects model (*b*=-0.02, 95% CI= - 0.05 to 0.01, *p* = 0.11), nor evidence of a time by congruency interaction in the interaction model (*b*=-0.01, 95% CI=-0.05 to 0.03, *p* = 0.62), with the congruent group increasing from 72% to 83% and the incongruent group from 70% to 79%. This suggests that, whilst participants did improve with training, this was not affected by whether participants were tested on the same or different stimuli to which they were trained on (Figure 5 and Supplementary Table S11).

**Figure 5.**
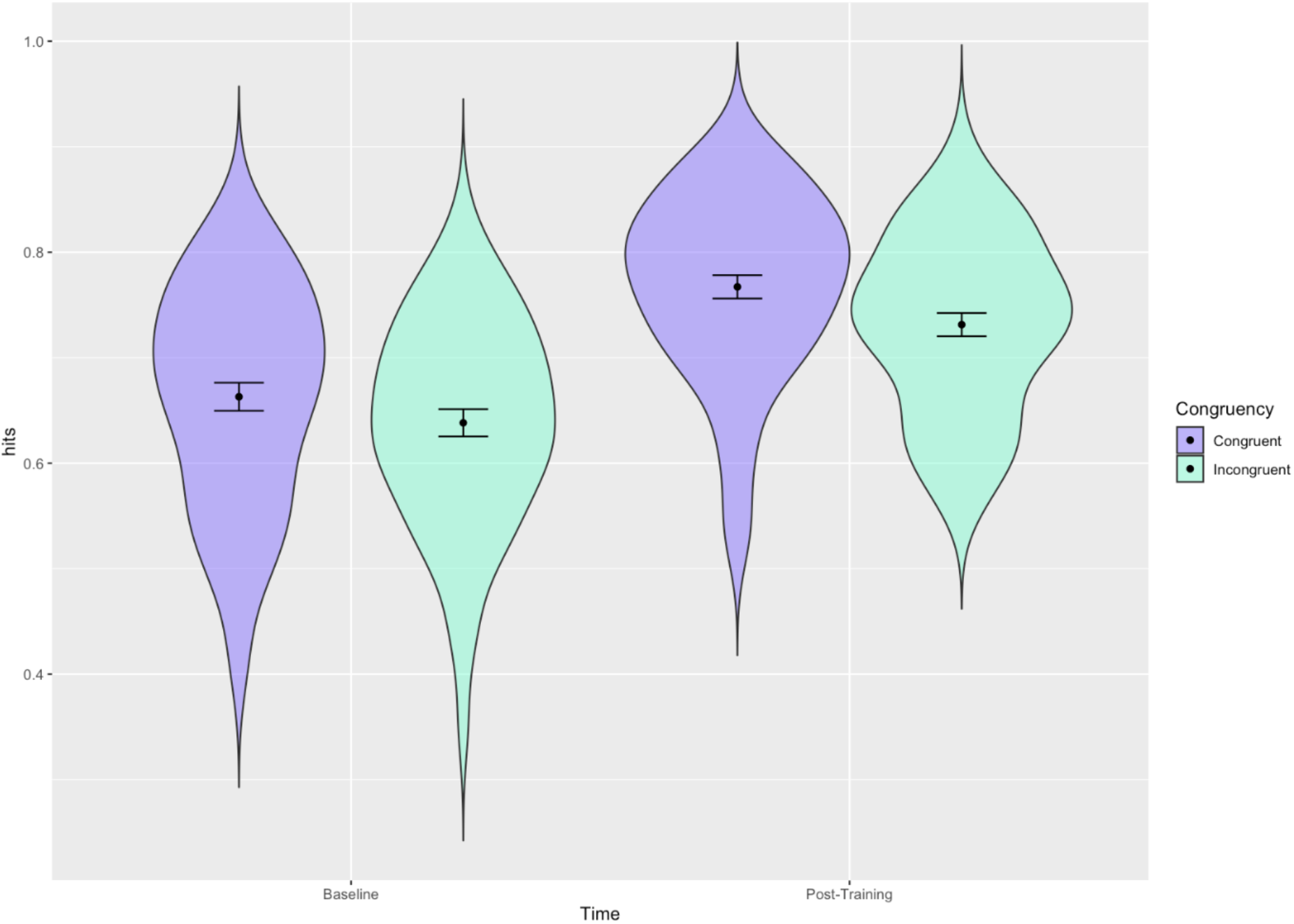
Distribution of participant’s total hits with estimates for the congruent and incongruent groups at baseline and post training in Study 3 (model including covariates). Error bars represent 95% confidence intervals: Distribution of participants scores (in terms of proportion of total correct hits) on the emotion recognition task, per congruency condition. Estimates for each condition before and after training and confidence intervals shown. Participants were either trained on stimuli that were the same (i.e., congruent) or that were different (i.e., incongruent) to the stimuli which they were tested on.

## 5. Discussion

Taken together, these studies provide evidence that 6AFC ERT improves emotion recognition accuracy in healthy volunteers. We demonstrate that it is possible to adapt an emotion recognition test into a training task, as has been done previously for emotional bias tests (Penton-Voak et al., 2012, 2013; Suddell et al., 2021). In our first study, using only four emotions, we did not find evidence of an improvement post-training in our active group compared to our sham group, although the direction of effect was as expected. However, we observed possible ceiling effects, as participants performed well at baseline, which likely reduced our ability to detect any improvement. In addition, any improvement may have been diluted by practice effects in the sham group. When the difficulty of the task was increased by adding two additional emotions, we found evidence for an improvement post-training in our active group compared to our sham group; thereby supporting the use of this training task as a potential intervention to improve emotion recognition. Such an intervention could be targeted at autistic individuals who experience difficulties with emotion recognition, for example. In addition, Study 3 suggested that this improvement in accuracy transfers to novel (non-trained) facial stimuli, suggesting that our results were not driven by repeat exposure to one set of stimuli. These findings are consistent with previous research supporting generalisability of facial emotion training (Dalili et al., 2016; Griffiths et al., 2015).

In Study 2 we additionally observed that improvements were higher for the scared and sad emotions. There was also a decrease in false alarms for surprised and sad, with overall improvements in the sensitivity score for sad, scared and surprised. However, we did not observe these effects across studies so further studies examining emotion specific hits, false alarms and sensitivity scores are needed to identify if any emotions in particular are impacted more by the training. Finally, we note that including the AQ-10 scores in Study 2 did not make a difference to our results, suggesting that participant’s level of autistic traits did not impact our training effect. However, as this was a sample of healthy volunteers, AQ-10 scores are likely to be lower than in autistic individuals. ERT may be more beneficial for autistic individuals where emotion recognition difficulties have been previously reported and baseline emotion recognition accuracy is likely to be lower (Law Smith et al., 2010; Lozier et al., 2014; Reed et al., 2021). Thus, it would be useful for future research to examine the effectiveness of ERT in an autistic sample and other populations, that may particularly stand to benefit from improved facial emotion recognition accuracy.

In addition, participants in our studies only completed one session of training, indicating a single session of training can provide some improvement in emotion recognition. However, it is plausible that multiple sessions of training would result in greater improvement in emotion recognition which is sustained over time. Therefore, assessing how this training impacts emotion recognition in a multi-session study would be helpful and it would also be useful to identify how many sessions of training would be optimal for delivery of this type of training.

### Limitations and future directions

There are some limitations to our studies that should be considered. First, while our studies were well-powered to detect effects, we did not find evidence for a training effect (i.e., greater improvement in total hits in the active group compared to sham) in Study 1 - we only observed a trend of improvement. However, as mentioned, this may have been due to ceiling effects leaving little room for improvement. These ceiling effects likely stemmed from our sample consisting of healthy adults with no known emotion recognition difficulties. These improvements were less than those for the active group, but may suggest there is a benefit from simply being exposed to the stimuli (i.e., in the baseline test). However, in Study 2 we did observe a training effect, likely due to the increased difficulty of the task. Therefore, future work with this training should include the six emotions used in Studies 2 and 3 and explore this training in populations with emotion recognition difficulties. Second, although Study 3 demonstrated generalisability to other faces, we have still only tested this for white male and female composite faces and thus generalisability beyond this is unknown. It would be useful to examine this further in future studies to assess generalisability to facial stimuli of different ethnicities and ages. Third, our study was conducted online which has some limitations, for example, it may be more difficult to be sure that participants actually pay attention to the study and are honest in their responses. However, there are many benefits to running studies online, such as the availability of a large sample pool living in different areas, so a broader sample of participants can be recruited.

## Conclusion

We find ERT improves emotion recognition accuracy in a sample of healthy volunteers, and an indication that training gains can transfer to novel faces (as we found no evidence of a difference between the congruent and incongruent conditions). This approach provides a strong basis for future studies around our training task, including testing this in autistic individuals and assessing the effectiveness of a multi-session approach.

## Data Availability

The data and analysis code that form the basis of the results presented here for all studies are available from the University of Bristol's Research Data Repository (http://data.bris.ac.uk/data/), DOI: https://doi.org/10.5523/bris.1df0stlnxblc72a13mfnsne3ew.

## Acknowledgements

We would like the thank all of the research participants that took part in our study.

## Author contributions

Conceptualization: ZER, SS, ASA; Methodology: ZER, SS, AE, ASA, ISPV; Formal Analysis: ZER, SS, AE, LT, ID; Resources: ZER, ASA; Data Curation: ZER; Writing— Original Draft: ZER, SS, LT, ID; Writing—Review and Editing: ZER, SS, AE, LT, ID, ISPV, CJ, MRM, ASA; Project Administration: ZER, ASA; Funding Acquisition: ZER, MRM, ASA.

