## Supplementary Materials for "Assessing the effectiveness of online emotion recognition training in healthy volunteers"

***Section 1. Ten-Item Autism Spectrum Quotient (AQ-10)***

We included questions from an abridged version of the Autism Spectrum Quotient, made up of 10 items (AQ-10) (Allison et al., 2012; Booth et al., 2013). Participants were asked whether descriptive statements related to them using the options “Definitely Agree”, “Slightly Agree”, “Slightly Disagree” or “Definitely Disagree”. The statements were:

- “I often notice small sounds when others do not”
- “I usually concentrate more on the whole picture, rather than the small details”
- “I find it easy to do more than one thing at once”
- “If there is an interruption, I can switch back to what I was doing very quickly”
- “I find it easy to ‘read between the lines’ when someone is talking to me”
- “I know how to tell if someone listening to me is getting bored”
- “When I’m reading a story I find it difficult to work out the characters’ intentions”
- “I like to collect information about categories of things”
- “I find it easy to work out what someone is thinking or feeling just by looking at their face”
- “I find it difficult to work out people’s intentions”.

***Section 2. Studies 1 to 3 skewness and kurtosis measures.***

Skewness and kurtosis measures for baseline and post-training emotion recognition hits across all three studies are shown in Supplementary Table S12. All values were within an acceptable range (Kim, 2013). Therefore, no transformations of these data were necessary.

**Supplementary Figure S1.** Histograms of the distribution of total hits as a proportion at baseline (a) and post-training (b) for study 1.

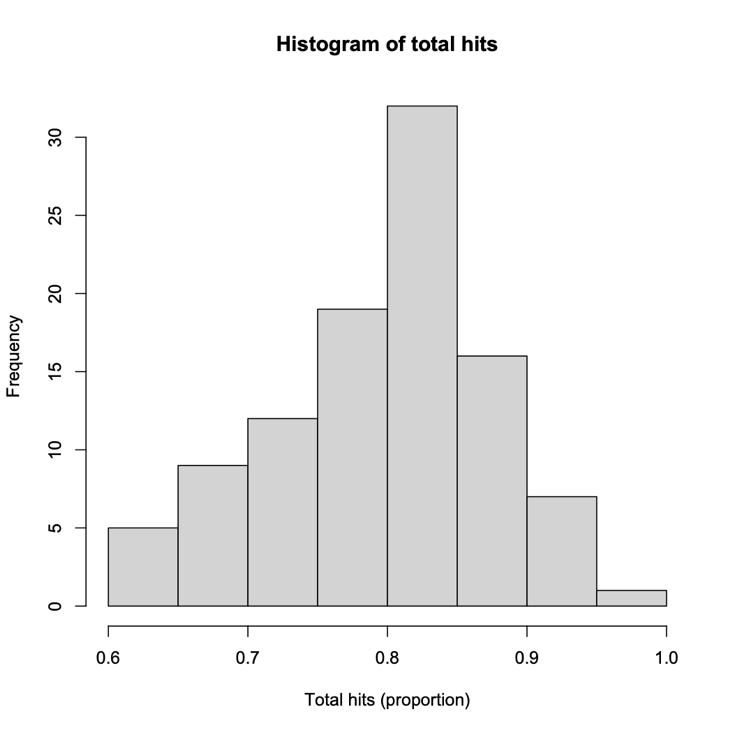

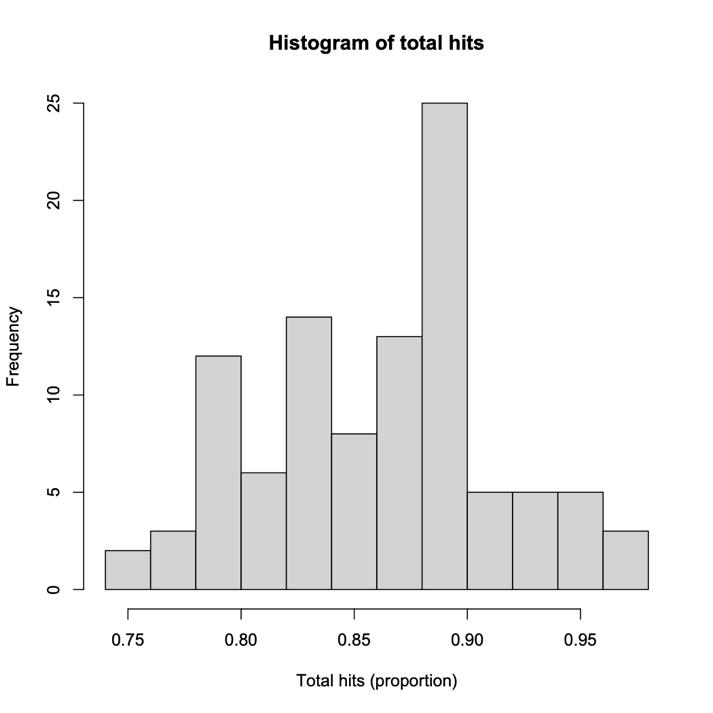

a)

b)

**Supplementary Figure S2.** Histograms of the distribution of total hits as a proportion at baseline (a) and post-training (b) for study 2.

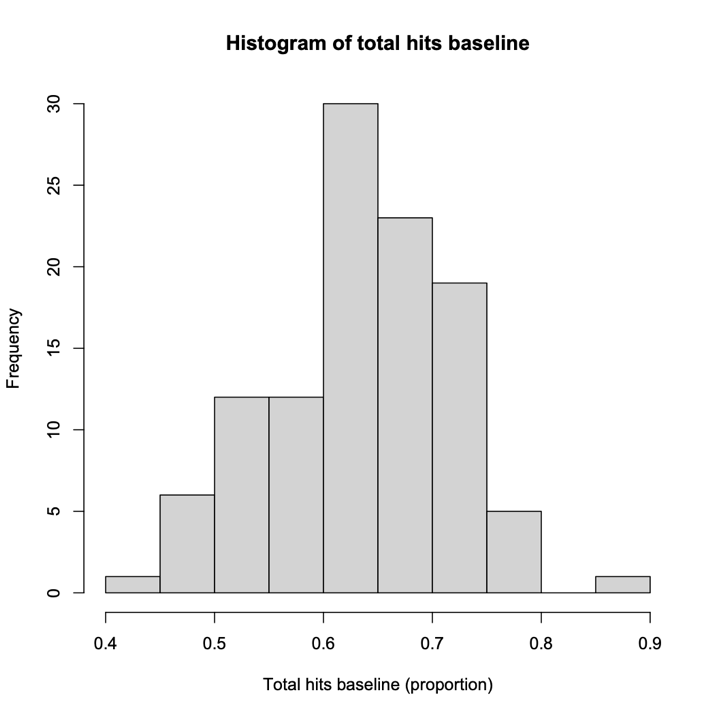

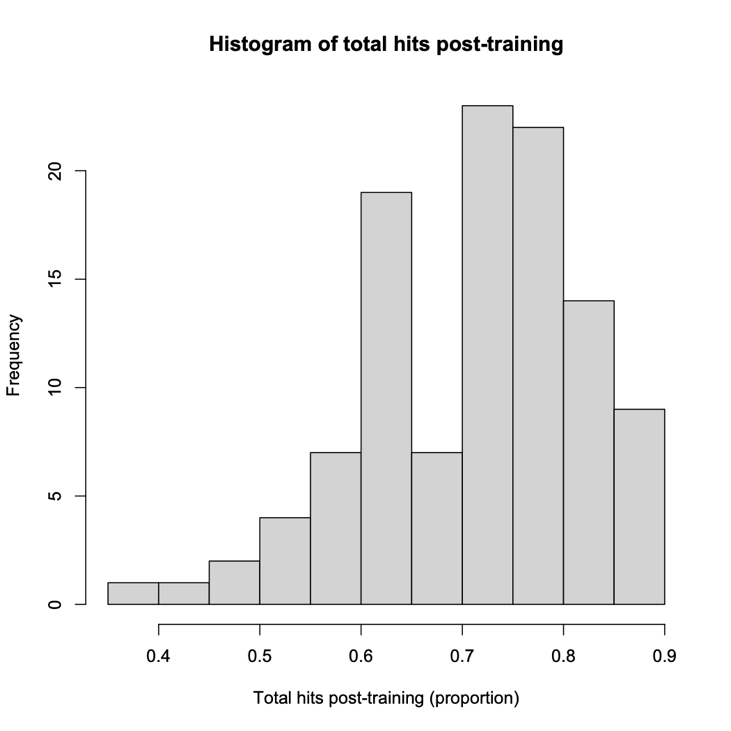

a)

b)

**Supplementary Figure S3.** Histograms of the distribution of total hits as a proportion at baseline (a) and post-training (b) for study 3.

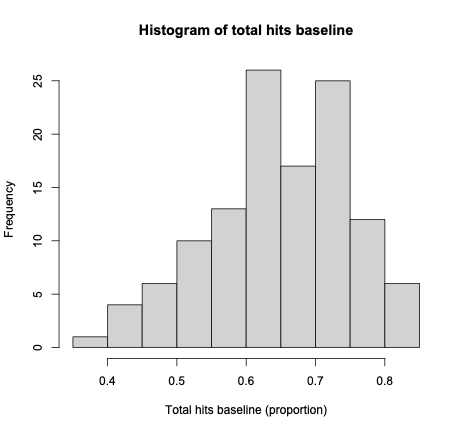

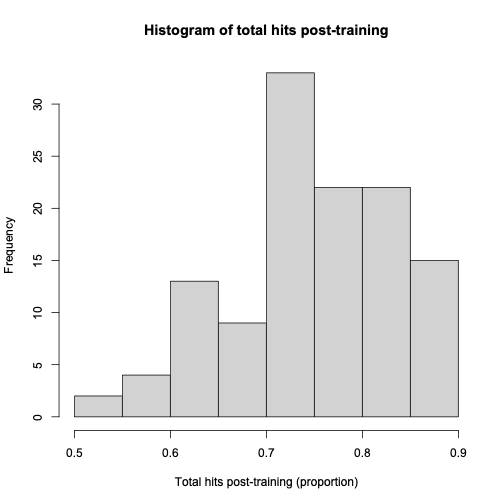

a)

b)

**Supplementary Table S1.** Study 1 results from the LME models for total hits including an interaction between time and group.

|  | Main effects model | | Interaction model (time*group) | |
| --- | --- | --- | --- | --- |
| *Fixed effects* | | | | |
| Predictors | ***b* (95% CI)** | **p-value** | ***b* (95% CI)** | **p-value** |
| Intercept | 0.79 (0.77 to 0.81) | 7.56x10^-163^ | 0.80 (0.78 to 0.82) | 2.25x10^-210^ |
| Time (post training) | 0.06 (0.04 to 0.08) | 1.90x10^-07^ | 0.05 (0.01 to 0.08) | 0.004 |
| Group (active) | 0.02 (0.002 to 0.05) | 0.03 | 0.01 (-0.02 to 0.04) | 0.46 |
| Time (post training) x group (active) |  |  | 0.02 (-0.02 to 0.07) | 0.27 |

*Results from the main effects and interaction LME models from study 1 (with an interaction of time and group) are presented here, where the beta estimate indicates the amount by which the proportion of total hits is increased for that predictor. CI= confidence interval. LME= Linear mixed effects.*

**Supplementary Table S2.** Study 1 results for the interaction between time (post-training) and group (active) from the LME models for emotion specific hits.

|  | **Angry** | | | | **Happy** | | | | **Sad** | | | | **Scared** | | | |
| --- | --- | --- | --- | --- | --- | --- | --- | --- | --- | --- | --- | --- | --- | --- | --- | --- |
|  | **Main effects model** | | **Interaction model (time*group)** | | **Main effects model** | | **Interaction model (time*group)** | | **Main effects model** | | **Interaction model (time*group)** | | **Main effects model** | | **Interaction model (time*group)** | |
| ***Fixed effects*** | | | | | | | | | | | | | | | | |
| **Predictors** | ***b* (95% CI)** | **p-value** | ***b* (95% CI)** | **p-value** | ***b* (95% CI)** | **p-value** | ***b* (95% CI)** | **p-value** | ***b* (95% CI)** | **p-value** | ***b* (95% CI)** | **p-value** | ***b* (95% CI)** | **p-value** | ***b* (95% CI)** | **p-value** |
| Intercept | 0.62 (0.59 to 0.66) | 4.75x10^-05^ | 0.64 (0.60 to 0.68) | 5.74x10^-72^ | 0.85 (0.82 to 0.89) | 5.18x10^-91^ | 0.86 (0.82 to 0.90) | 2.61x10^-96^ | 0.75 (0.71 to 0.78) | 1.07x10^-83^ | 0.75 (0.72 to 0.79) | 2.41x10^-90^ | 0.95 (0.93 to 0.97) | 5.84x10^-140^ | 0.94 (0.93 to 0.96) | 1.72x10^-156^ |
| Time (post training) | 0.12 (0.09 to 0.15) | 5.77x10^-12^ | 0.09 (0.04 to 0.13) | 1.53x10^-04^ | 0.04 (0.01 to 0.08) | 0.01 | 0.03 (-0.02 to 0.08) | 0.19 | 0.05 (0.02 to 0.08) | 4.39x10^-04^ | 0.04 (-0.004 to 0.08) | 0.08 | 0.01 (-0.001 to 0.03) | 0.07 | 0.03 (0.004 to 0.05) | 0.02 |
| Group (active) | 0.06 (0.01 to 0.11) | 0.01 | 0.03 (-0.03 to 0.08) | 0.37 | -0.03 (-0.07 to 0.01) | 0.19 | -0.04 (-0.10 to 0.02) | 0.16 | 0.06 (0.01 to 0.10) | 0.01 | 0.04 (-0.01 to 0.09) | 0.11` | 0.01 (-0.01 to 0.03) | 0.45 | 0.02 (-001 to 0.05) | 0.14 |
| Time (post training) x group (active) |  |  | 0.07 (0.01 to 0.13) | 0.03 |  |  | 0.02 (-0.05 to 0.09) | 0.54 |  |  | 0.03 (-0.03 to 0.09) | 0.30 |  |  | -0.02 (-0.05 to 0.01) | 0.13 |

*Results from the main effects and interaction LME models from study 1 for hits (with an interaction of time and group) are presented here, where the beta estimate indicates the amount by which the proportion of total hits is increased for that predictor. CI= confidence interval. LME= Linear mixed effects.*

**Supplementary Table S3.** Study 1 results for the interaction between time (post-training) and group (active) from the LME models for emotion specific false alarms.

|  | **Angry** | | | | **Happy** | | | | **Sad** | | | | **Scared** | | | | |
| --- | --- | --- | --- | --- | --- | --- | --- | --- | --- | --- | --- | --- | --- | --- | --- | --- | --- |
|  | **Main effects model** | | **Interaction model (time*group)** | | **Main effects model** | | **Interaction model (time*group)** | | **Main effects model** | | **Interaction model (time*group)** | | **Main effects model** | | | **Interaction model (time*group)** | |
| ***Fixed effects*** | | | | | | | | | | | | | | | | | |
| **Predictors** | ***b* (95% CI)** | **p-value** | ***b* (95% CI)** | **p-value** | ***b* (95% CI)** | **p-value** | ***b* (95% CI)** | **p-value** | ***b* (95% CI)** | **p-value** | ***b* (95% CI)** | **p-value** | ***b* (95% CI)** | **p-value** | ***b* (95% CI)** | | **p-value** |
| Intercept | 0.02 (0.01 to 0.03) | 1.50x10^-06^ | 0.02 (0.01 to 0.03) | 5.76x10^-07^ | 0.11 (0.09 to 0.13) | 4.64x10^-24^ | 0.10 (0.08 to 0.12) | 1.85x10^-19^ | 0.10 (0.08 to 0.12) | 1.62x10^-21^ | 0.10 (0.08 to 0.12) | 1.91x10^-19^ | 0.05 (0.04 to 0.06) | 1.83x10^-12^ | 0.05 (0.03 to 0.06) | | 7.92x10^-11^ |
| Time (post training) | -0.004 (-0.01 to 0.003) | 0.26 | -0.01 (-0.02 to 0.0006) | 0.07 | -0.03 (-0.04 to -0.01) | 0.001 | -0.01 (-0.03 to 0.02) | 0.52 | -0.02 (-0.04 to -0.004) | 0.02 | -0.02 (-0.05 to 0.001) | 0.07 | -0.03 (-0.04 to -0.02) | 3.63x10^-07^ | -0.02 (-0.04 to -0.01) | | 0.001 |
| Group (active) | 0.001 (-0.01 to 0.01) | 0.81 | 0.009 (-0.01 to 0.01) | 0.56 | -0.03 (-0.05 to -0.01) | 0.02 | -0.01 (-0.04 to 0.02) | 0.57 | -0.01 (-0.03 to 0.02) | 0.60 | -0.01 (-0.03 to 0.02) | 0.57 | 0.0006 (-0.01 to 0.02) | 0.94 | 0.004 (-0.01 to 0.02) | | 0.68 |
| Time (post training) x group (active) |  |  | 0.01 (-0.003 to 0.02) | 0.15 |  |  | -0.04 (-0.07 to -0.01) | 0.02 |  |  | 0.004 (-0.03 to 0.04) | 0.80 |  |  | -0.01 (-0.03 to 0.01) | | 0.51 |

*Results from the main effects and interaction LME models from study 1 for false alarms (with an interaction of time and group) are presented here, where the beta estimate indicates the amount by which the proportion of false alarms score is increased for that predictor. CI= confidence interval. LME= Linear mixed effects.*

**Supplementary Table S4.** Study 1 results for the interaction between time (post-training) and group (active) from the LME models for emotion specific sensitivity scores.

|  | **Angry** | | | | | **Happy** | | | | | | **Sad** | | | | | | **Scared** | | | | |
| --- | --- | --- | --- | --- | --- | --- | --- | --- | --- | --- | --- | --- | --- | --- | --- | --- | --- | --- | --- | --- | --- | --- |
|  | **Main effects model** | | **Interaction model (time*group)** | | | **Main effects model** | | | **Interaction model (time*group)** | | | **Main effects model** | | | **Interaction model (time*group)** | | | **Main effects model** | | | **Interaction model (time*group)** | |
| ***Fixed effects*** | | | | | | | | | | | | | | | | | | | | | | |
| **Predictors** | ***b* (95% CI)** | **p-value** | | ***b* (95% CI)** | **p-value** | | ***b* (95% CI)** | **p-value** | | ***b* (95% CI)** | **p-value** | | ***b* (95% CI)** | **p-value** | | ***b* (95% CI)** | **p-value** | | ***b* (95% CI)** | **p-value** | ***b* (95% CI)** | **p-value** |
| Intercept | 0.90 (0.89 to 0.91) | 3.25x10^-165^ | | 0.90 (0.89 to 0.91) | 4.57x10^-189^ | | 0.93 (0.92 to 0.94) | 7.51x10^-192^ | | 0.93 (0.92 to 0.94) | 1.17x10^-122^ | | 0.90 (0.89 to 0.91) | 5.17x10^-174^ | | 0.90 (0.89 to 0.91) | 1.02x10^-197^ | | 0.97 (0.97 to 0.98) | 1.71x10^-200^ | 0.97 (0.97 to 0.98) | 1.54x10^-232^ |
| Time (post training) | 0.03 (0.03 to 0.04) | 2.37x10^-12^ | | 0.03 (0.02 to 0.04) | 1.68x10^-05^ | | 0.02 (0.01 to 0.03) | 2.92x10^-05^ | | 0.01 (-0.001 to 0.03) | 0.08 | | 0.02 (0.01 to 0.03) | 1.55x10^-06^ | | 0.02 (0.01 to 0.04) | 0.002 | | 0.01 (0.01 to 0.02) | 7.91x10^-05^ | 0.01 (0.01 to 0.02) | 0.001 |
| Group (active) | 0.02 (0.003 to 0.03) | 0.02 | | 0.01 (-0.01 to 0.02) | 0.22 | | -0.002 (-0.01 to 0.01) | 0.79 | | -0.01 (-0.03 to 0.004) | 0.17 | | 0.02 (0.01 to 0.03) | 0.006 | | 0.02 (-0.00008 to 0.03) | 0.05 | | 0.001 (-0.01 to 0.01) | 0.74 | 0.003 (-0.01 to 0.01) | 0.48 |
| Time (post training) x group (active) |  |  | | 0.01 (-0.004 to 0.03) | 0.14 | |  |  | | 0.02 (-0.001 to 0.04) | 0.07 | |  |  | | 0.005 (-0.01 to 0.02) | 0.64 | |  |  | -0.004 (-0.01 to 0.01) | 0.44 |

*Results from the main effects and interaction LME models from study 1 for sensitivity scores (with an interaction of time and group) are presented here, where the beta estimate indicates the amount by which the sensitivity score is increased for that predictor. CI= confidence interval. LME= Linear mixed effects.*

**Supplementary Table S5.** Study 1 results for t-tests of group differences in subjective ratings of the study and PANAS positive and negative scores.

|  | **Active group mean (SD)** | **Sham group mean (SD)** | **T-test p-value** |
| --- | --- | --- | --- |
| **Fatiguing** | 42 (29) | 36 (25) | 0.33 |
| **Challenging** | 63 (23) | 54 (28) | 0.09 |
| **Interesting** | 67 (23) | 69 (25) | 0.54 |
| **PANAS positive** | 27 (8) | 30 (8) | 0.07 |
| **PANAS negative** | 14 (5) | 13 (2) | 0.15 |

*The mean and standard deviations (SD) for each item per group are presented here along with the p-value from t-tests conducted. We are comparing these subjective ratings across groups as the tasks completed were different and we might expect there to be differences in how participants found the tasks. We also wanted to see whether there was any indication of differences in PANAS for those that received the emotion recognition training compared to those that did not. PANAS=* *Positive and Negative Affect Schedule.*

**Supplementary Table S6.** Study 2 results from the LME models for total hits including an interaction between time and group.

|  | **Unadjusted** | | | | **Adjusted for age, gender and education** | | | | **Adjusted additionally for AQ-10** | | | |
| --- | --- | --- | --- | --- | --- | --- | --- | --- | --- | --- | --- | --- |
|  | **Main effects model** | | **Interaction model (time*group)** | | **Main effects model** | | **Interaction model (time*group)** | | **Main effects model** | | **Interaction model (time*group)** | |
| ***Fixed effects*** | | | | | | | | | | | | |
| **Predictors** | ***b* (95% CI)** | **p-value** | ***b* (95% CI)** | **p-value** | ***b* (95% CI)** | **p-value** | ***b* (95% CI)** | **p-value** | ***b* (95% CI)** | **p-value** | ***b* (95% CI)** | **p-value** |
| Intercept | 0.62 (0.60 to 0.64) | 3.19x10-107 | 0.64 (0.61 to 0.66 | 1.10x10-128 | 0.68 (0.62 to 0.74) | 1.55x10-43 | 0.70 (0.64 to 0.76) | 1.71x10-45 | 0.68 (0.61 to 0.75) | 8.81x10-37 | 0.70 (0.63 to 0.77) | 3.56x10-38 |
| Time (post training) | 0.07 (0.05 to 0.09) | 3.57x10-09 | 0.03 (0.002 to 0.07) | 0.04 | 0.07 (0.05 to 0.09) | 3.57x10-09 | 0.03 (0.002 to 0.07) | 0.04 | 0.07 (0.05 to 0.09) | 3.57x10-09 | 0.03 (0.002 to 0.07) | 0.04 |
| Group (active) | 0.04 (0.01 to 0.07) | 0.006 | 0.01 (-0.03 to 0.04) | 0.73 | 0.04 (0.01 to 0.07) | 0.005 | 0.01 (-0.03 to 0.05) | 0.68 | 0.04 (0.01 to 0.07) | 0.005 | 0.01 (-0.03 to 0.05) | 0.68 |
| Time (post training) x group (active) |  |  | 0.07 (0.03 to 0.12) | 0.002 |  |  | 0.07 (0.03 to 0.12) | 0.002 |  |  | 0.07 (0.03 to 0.12) | 0.002 |

*Results from the unadjusted and adjusted main effects and interaction LME models from study 2 (with an interaction of time and group) are presented here, where the beta estimate indicates the amount by which the proportion of total hits is increased for that predictor. CI= confidence interval. LME= Linear mixed effects, AQ-10= 10-Item Autism Spectrum Quotient.*

**Supplementary Table S7.** Study 2 results for the interaction between time (post-training) and group (active) from the LME models for emotion specific hits.

|  | **Angry** | | | | **Happy** | | | | | **Sad** | | | | **Scared** | | | | **Surprised** | | | | **Disgust** | | | |
| --- | --- | --- | --- | --- | --- | --- | --- | --- | --- | --- | --- | --- | --- | --- | --- | --- | --- | --- | --- | --- | --- | --- | --- | --- | --- |
|  | **Main effects model** | | **Interaction model (time*group)** | | **Main effects model** | | **Interaction model (time*group)** | | | **Main effects model** | | **Interaction model (time*group)** | | **Main effects model** | | **Interaction model (time*group)** | | **Main effects model** | | **Interaction model (time*group)** | | **Main effects model** | | **Interaction model (time*group)** | |
| ***Fixed effects*** | | | | | | | | | | | | | | | | | | | | | | | | | |
| **Predictors** | ***b* (95% CI)** | **p-value** | ***b* (95% CI)** | **p-value** | ***b* (95% CI)** | **p-value** | | ***b* (95% CI)** | **p-value** | ***b* (95% CI)** | **p-value** | ***b* (95% CI)** | **p-value** | ***b* (95% CI)** | **p-value** | ***b* (95% CI)** | **p-value** | ***b* (95% CI)** | **p-value** | ***b* (95% CI)** | **p-value** | ***b* (95% CI)** | **p-value** | ***b* (95% CI)** | **p-value** |
| Intercept | 0.53 (0.43 to 0.63) | 7.46x10^-18^ | 0.54 (0.43 to 0.64) | 3.19x10^-18^ | 0.65 (0.51 to 0.78) | 2.25x10^-16^ | | 0.66 (0.53 to 0.80) | 6.75x10^-17^ | 0.74 (0.64 to 0.85) | 3.78x10^-27^ | 0.76 (0.66 to 0.86) | 6.13x10^-28^ | 0.58 (0.40 to 0.75) | 2.41x10^-09^ | 0.64 (0.47 to 0.82) | 7.88x10^-11^ | 0.76 (0.68 to 0.84) | 1.60x10^-36^ | 0.76 (0.68 to 0.84) | 5.82x10^-37^ | 0.84 (0.73 to 0.95) | 1.05x10^-28^ | 0.84 (0.73 to 0.95) | 8.84x10^-29^ |
| Time (post training) | 0.10 (0.07 to 0.14) | 3.50x10^-08^ | 0.08 (0.03 to 0.13) | 0.001 | 0.11 (0.06 to 0.15) | 3.75x10^-06^ | | 0.07 (0.01 to 0.13) | 0.02 | 0.02 (-0.01 to 0.05) | 0.26 | -0.01 (-0.06 to 0.03) | 0.57 | 0.16 (0.11 to 0.21) | 3.45x10^-08^ | 0.03 (-0.04 to 0.10) | 0.38 | -0.01 (-0.04 to 0.03) | 0.72 | Add (-0.05 to 0.04) | 0.84 | 0.04 (ADD to 0.08) | 0.08 | 0.04 (-0.02 to 0.10) | 0.17 |
| Group (active) | 0.04 (-0.01 to 0.10) | 0.09 | 0.02 (-0.04 to 0.09) | 0.44 | 0.08 (0.02 to 0.15) | 0.02 | | 0.05 (-0.03 to 0.13) | 0.24 | -0.01 (-0.06 to 0.05) | 0.83 | -0.04 (-0.10 to 0.02) | 0.22 | 0.14 (0.05 to 0.23) | 0.003 | 0.01 (-0.09 to 0.11) | 0.84 | -0.01 (-0.05 to 0.03) | 0.50 | -0.01 (-0.06 to 0.04) | 0.63 | 0.02 (-0.04 to 0.07) | 0.60 | 0.02 (-0.05 to 0.09) | 0.61 |
| Time (post training) x group (active) |  |  | 0.04 (-0.03 to 0.11) | 0.23 |  |  | | 0.07 (-0.01 to 0.16) | 0.10 |  |  | 0.07 (-0.0002 to 0.13) | 0.05 |  |  | 0.26 (0.17 to 0.36) | 2.47x10^-07^ |  |  | -0.002 (-0.07 to 0.06) | 0.94 |  |  | -0.01 (-0.09 to 0.08) | 0.89 |

*Results from the adjusted (for age, gender and education level) main effects and interaction LME models from study 2 for hits (with an interaction of time and group) are presented here, where the beta estimate indicates the amount by which the proportion of total hits is increased for that predictor. CI= confidence interval. LME= Linear mixed effects, Obs=Number of observations*

**Supplementary Table S8.** Study 2 results for the interaction between time (post-training) and group (active) from the LME models for emotion specific false alarms.

|  | **Angry** | | | | **Happy** | | | | | **Sad** | | | | **Scared** | | | | **Surprised** | | | | **Disgust** | | | |
| --- | --- | --- | --- | --- | --- | --- | --- | --- | --- | --- | --- | --- | --- | --- | --- | --- | --- | --- | --- | --- | --- | --- | --- | --- | --- |
|  | **Main effects model** | | **Interaction model (time*group)** | | **Main effects model** | | **Interaction model (time*group)** | | | **Main effects model** | | **Interaction model (time*group)** | | **Main effects model** | | **Interaction model (time*group)** | | **Main effects model** | | **Interaction model (time*group)** | | **Main effects model** | | **Interaction model (time*group)** | |
| ***Fixed effects*** | | | | | | | | | | | | | | | | | | | | | | | | | |
| **Predictors** | ***b* (95% CI)** | **p-value** | ***b* (95% CI)** | **p-value** | ***b* (95% CI)** | **p-value** | | ***b* (95% CI)** | **p-value** | ***b* (95% CI)** | **p-value** | ***b* (95% CI)** | **p-value** | ***b* (95% CI)** | **p-value** | ***b* (95% CI)** | **p-value** | ***b* (95% CI)** | **p-value** | ***b* (95% CI)** | **p-value** | ***b* (95% CI)** | **p-value** | ***b* (95% CI)** | **p-value** |
| Intercept | 0.01 (-0.01 to 0.03) | 0.18 | 0.01 (-0.01 to 0.03) | 0.18 | 0.02 (-0.02 to 0.05) | 0.36 | | 0.01 (-0.02 to 0.05) | 0.43 | 0.07 (0.04 to 0.11) | 1.38x10-04 | 0.07 (0.03 to 0.11) | 3.60x10-04 | 0.05 (0.02 to 0.08) | 0.002 | 0.05 (0.0 2 to 0.08) | 0.002 | 0.14 (0.09 to 0.19) | 1.09x10-07 | 0.13 (0.08 to 0.18) | 1.01x10-06 | 0.09 (0.05 to 0.13) | 2.93x10-05 | 0.09 (0.05 to 0.13) | 5.51x10-05 |
| Time (post training) | -0.003 (-0.01 to 0.004) | 0.38 | -0.004 (-0.01 to 0.01) | 0.52 | 0.01 (0.004 to 0.02) | 0.007 | | 0.02 (ADD to 0.03) | 0.01 | -0.02 (-0.03 to -0.01) | 8.87x10-06 | -0.02 (-0.03 to -0.0009) | 0.04 | -0.01 (-0.02 to -0.002) | 0.02 | -0.01 (-0.02 to 0.004) | 0.15 | -0.04 (-0.06 to -0.03) | 7.38x10-10 | -0.02 (-0.04 to ADD) | 0.02 | -0.02 (-0.03 to -0.002) | 0.02 | -0.01 (-0.03 to 0.01) | 0.27 |
| Group (active) | 0.00007 (-0.01 to 0.01) | 0.99 | -0.0001 (-0.01 to 0.01) | 0.98 | 0.01 (-0.01 to 0.02) | 0.58 | | 0.01 (-0.01 to 0.03) | 0.39 | -0.003 (-0.02 to 0.02) | 0.75 | 0.01 (-0.02 to 0.03) | 0.58 | -0.01 (-0.02 to 0.01) | 0.25 | -0.01 (-0.03 to 0.01) | 0.42 | -0.03 (-0.06 to -0.01) | 0.02 | -0.01 (-0.04 to 0.02) | 0.62 | -0.02 (-0.04 to 0.01) | 0.15 | -0.01 (-0.03 to 0.01) | 0.41 |
| Time (post training) x group (active) |  |  | 0.0004 (-0.02 to 0.02) | 0.96 |  |  | | -0.01 (-0.03 to 0.01) | 0.45 |  |  | -0.02 (-0.04 to 0.002) | 0.07 |  |  | 0.003 (-0.02 to 0.02) | 0.77 |  |  | -0.05 (-0.07 to -0.02) | 0.0001 |  |  | -0.01 (-0.04 to 0.02) | 0.48 |

*Results from the adjusted (for age, gender and education level) main effects and interaction LME models from study 2 for false alarms (with an interaction of time and group) are presented here, where the beta estimate indicates the amount by which the proportion of false alarms score is increased for that predictor. CI= confidence interval. LME= Linear mixed effects, Obs=Number of observations*

**Supplementary Table S9.** Study 2 results for the interaction between time (post-training) and group (active) from the LME models for emotion specific sensitivity scores.

|  | **Angry** | | | | **Happy** | | | | | **Sad** | | | | **Scared** | | | | **Surprised** | | | | **Disgust** | | | |
| --- | --- | --- | --- | --- | --- | --- | --- | --- | --- | --- | --- | --- | --- | --- | --- | --- | --- | --- | --- | --- | --- | --- | --- | --- | --- |
|  | **Main effects model** | | **Interaction model (time*group)** | | **Main effects model** | | **Interaction model (time*group)** | | | **Main effects model** | | **Interaction model (time*group)** | | **Main effects model** | | **Interaction model (time*group)** | | **Main effects model** | | **Interaction model (time*group)** | | **Main effects model** | | **Interaction model (time*group)** | |
| ***Fixed effects*** | | | | | | | | | | | | | | | | | | | | | | | | | |
| **Predictors** | ***b* (95% CI)** | **p-value** | ***b* (95% CI)** | **p-value** | ***b* (95% CI)** | **p-value** | | ***b* (95% CI)** | **p-value** | ***b* (95% CI)** | **p-value** | ***b* (95% CI)** | **p-value** | ***b* (95% CI)** | **p-value** | ***b* (95% CI)** | **p-value** | ***b* (95% CI)** | **p-value** | ***b* (95% CI)** | **p-value** | ***b* (95% CI)** | **p-value** | ***b* (95% CI)** | **p-value** |
| Intercept | 0.88 (0.85 to 0.90) | 4.58x10^-89^ | 0.88 (0.85 to 0.91) | 1.99x10^-93^ | 0.90 (0.86 to 0.94) | 7.94x10^-73^ | | 0.91 (0.87 to 0.95) | 5.93x10^-76^ | 0.90 (0.87 to 0.94) | 4.90x10^-75^ | 0.91 (0.88 to 0.95) | 5.62x10^-78^ | 0.87 (0.75 to 0.98) | 5.85x10^-28^ | 0.90 (0.79 to 1.02) | 1.43x10^-29^ | 0.87 (0.83 to 0.92) | 2.77x10^-68^ | 0.88 (0.84 to 0.92) | 2.64x10^-70^ | 0.94 (0.90 to 0.98) | 9.52x10^-73^ | 0.94 (0.90 to 0.98) | 2.72x10^-75^ |
| Time (post training) | 0.03 (0.02 to 0.04) | 4.86x10^-07^ | 0.03 (0.01 to 0.04) | 0.001 | 0.03 (0.01 to 0.04) | 0.001 | | 0.02 (-0.004 to 0.04) | 0.10 | 0.02 (0.004 to 0.03) | 0.01 | 0.01 (-0.02 to 0.02) | 0.94 | 0.05 (0.01 to 0.09) | 0.01 | -0.01 (-0.07 to 0.04) | 0.57 | 0.02 (0.003 to 0.03) | 0.02 | 0.01 (-0.01 to 0.02) | 0.61 | 0.02 (0.004 to 0.04) | 0.02 | 0.02 (-0.009 to 0.04) | 0.08 |
| Group (active) | 0.01 (-0.003 to 0.02) | 0.14 | 0.01 (-0.01 to 0.02) | 0.46 | 0.02 (0.004 to 0.04) | 0.02 | | 0.01 (-0.01 to 0.04) | 0.27 | -0.01 (-0.03 to 0.01) | 0.52 | -0.02 (-0.05 to -0.0007) | 0.05 | 0.09 (0.03 to 0.15) | 0.004 | 0.02 (-0.05 to 0.09) | 0.49 | 0.004 (-0.02 to 0.03) | 0.70 | -0.01 (-0.03 to 0.02) | 0.55 | 0.01 (-0.01 to 0.03) | 0.39 | 0.01 (-0.02 to 0.04) | 0.46 |
| Time (post training) x group (active) |  |  | 0.01 (-0.02 to 0.03) | 0.51 |  |  | | 0.02 (-0.01 to 0.05) | 0.26 |  |  | 0.04 (0.01 to 0.07) | 0.01 |  |  | 0.13 (0.05 to 0.20) | 0.001 |  |  | 0.02 (-0.003 to 0.05) | 0.09 |  | 0.14 | -0.001 (-0.03 to 0.03) | 0.93 |
| Marginal R2/ Conditional R2 | 0.14/0.29 | | 0.14/0.29 | | 0.10/0.29 | | 0.10/0.29 | | | 0.04/0.28 | | 0.06/0.31 | | 0.12/0.48 | | 0.14/0.53 | | 0.05/0.42 | | 0.06/0.43 | | 0.07/0.31 | | 0.07/0.30 | |

*Results from the adjusted (for age, gender and education level) main effects and interaction LME models from study 2 for sensitivity scores (with an interaction of time and group) are presented here, where the beta estimate indicates the amount by which the sensitivity score is increased for that predictor. CI= confidence interval. LME= Linear mixed effects, Obs=Number of observations*

**Supplementary Table S10.** Study 2 results for t-tests of group differences in subjective ratings of the study.

|  | **Active group mean (SD)** | **Control group mean (SD)** | **T-test p-value** |
| --- | --- | --- | --- |
| **Fatiguing** | 43 (27) | 39 (30) | 0.52 |
| **Challenging** | 69 (22) | 60 (30) | 0.10 |
| **Interesting** | 71 (23) | 65 (24) | 0.16 |

*The mean and standard deviations (SD) for each item per group are presented here along with the p-value from t-tests conducted.*

**Supplementary Table S11.** Study 3 results from the LME models for total hits including an interaction between time and congruency.

|  | **Unadjusted** | | | | **Adjusted for age, gender and education** | | | |
| --- | --- | --- | --- | --- | --- | --- | --- | --- |
|  | **Main effects model** | | **Interaction model (time*group)** | | **Main effects model** | | **Interaction model (time*group)** | |
| ***Fixed effects*** | | | | | | | | |
| **Predictors** | ***b* (95% CI)** | **p-value** | ***b* (95% CI)** | **p-value** | ***b* (95% CI)** | **p-value** | ***b* (95% CI)** | **p-value** |
| Intercept | 0.67 (0.64 to 0.69) | 2.27x10^-123^ | 0.66 (0.64 to 0.69) | 7.84x10^-145^ | 0.72 (0.67 to 0.78) | 6.34x10^-50^ | 0.72 (0.66 to 0.78) | 1.05x10^-50^ |
| Time (post training) | 0.10 (0.08 to 0.12) | 2.90x10^-18^ | 0.10 (0.07 to 0.13) | 3.05x10^-11^ | 0.10 (0.08 to 0.12) | 2.90x10^-18^ | 0.10 (0.07 to 0.13) | 3.05x10-11 |
| Congruency (incongruent) | -0.03 (-0.06 to -0.001) | 0.04 | -0.02 (-0.06 to 0.01) | 0.19 | -0.02 (-0.05 to 0.01) | 0.11 | -0.02 (-0.06 to 0.02) | 0.32 |
| Time (post training) x congruency (active) |  |  | -0.01 (-0.05 to 0.03) | 0.62 |  |  | -0.01 (-0.05 to 0.03) | 0.62 |

*Results from the unadjusted and adjusted (for age, gender and education level) main effects and interaction LME models from study 3 (with an interaction of time and congruency) are presented here, where the beta estimate indicates the amount by which the proportion of total hits is increased for that predictor. CI= confidence interval. LME= Linear mixed effects.*

**Supplementary Table S12.** Skewness and kurtosis measures across studies for emotion recognition hits.

|  | **Baseline** | **Post-training** |
| --- | --- | --- |
| ***Study 1*** | | |
| **Skewness** | -0.48 | -0.06 |
| **Kurtosis** | 0.07 | 0.65 |
| ***Study 2*** | | |
| **Skewness** | -0.14 | -0.63 |
| **Kurtosis** | -0.26 | -0.09 |
| ***Study 3*** | | |
| **Skewness** | -0.40 | -0.48 |
| **Kurtosis** | -0.28 | -0.32 |
